# Coronavirus Disease 2019 (COVID-19) Dynamics Considering the Influence of Hospital Infrastructure

**DOI:** 10.1101/2020.06.03.20121608

**Authors:** Pedro M.C.L. Pacheco, Marcelo A. Savi, Pedro V. Savi

## Abstract

This paper deals with the mathematical modeling and numerical simulations related to the coronavirus dynamics. A description is developed based on the framework of susceptible-exposed-infectious-removed model. Removed population is split into recovered and death populations allowing a better comprehension of real situations. Besides, total population is reduced based on the number of deaths. Hospital infrastructure is also included into the mathematical description allowing the consideration of collapse scenarios. Initially, a model verification is carried out calibrating system parameters with data from China outbreak that is considered a benchmark due the availability of data for the entire cycle. Afterward, numerical simulations are performed to analyze COVID-19 dynamics in Brazil. Results show several scenarios showing the importance of social isolation. System dynamics has a strong sensitivity to transmission rate showing the importance of numerical simulations to guide public health decision strategies.

## 1. INTRODUCTION

Coronavirus disease 2019 (COVID-19) was discovered in 2019, becoming a pandemic that is promoting a dramatic reaction all over the world. Several uncertainties are associated with all the aspects of this disease including clinical evolution and contamination processes. Mathematical modeling is an interesting approach that can allow the evaluation of different scenarios, furnishing information for a proper support for health system decisions. Nonlinear dynamics of biological and biomedical systems is the objective of several researches that can be based on mathematical modeling or time series analysis (Savi, 2005). In particular, coronavirus spread can be described by a mathematical model that allows the nonlinear dynamics analysis, representing different populations related to the phenomenon.

Different kinds of models can be employed for the COVID-19 dynamics. Rihan *et al*. (2018) described the dynamics of coronavirus infection in human, establishing interaction among human cells and the virus. Chen *et al*. (2020) developed a mathematical model for calculating the transmissibility of the virus considering a simplified version of the bats-hosts-reservoir-people transmission model, defined as a reservoir-people model. Li *et al*. (2020b) estimated characteristics of the epidemiologic time distribution, exploiting some pattern trends of transmission propagation. Riou & Althaus (2020) exploited the pattern of human-to-human transmission of novel coronavirus in Wuhan, China. Two key parameters are considered: basic reproduction number that defines the infectious propagation; and the individual variation in the number of secondary cases. Uncertainty quantification tools were employed to define the transmission patterns.

Susceptible-exposed-infectious-removed (SEIR) models are an interesting approach to deal with the mathematical modeling of coronavirus transmission. Wu *et al*. (2020) investigated Wuhan – China case, evaluating domestic and international spread outbreak. Lin *et al*. (2020) proposed a model considering individual reaction, governmental action and emigration. The model is based on the original work of He *et al*. (2013) that proposed a model to describe the 1918 influenza.

COVID-19 scenarios all over the world are becoming dramatic due to the absence of effective drugs and/or vaccines. Since it is possible that this general scenario is persisting for an unknown period of time, it is required that governments need to implement alternative strategies, known as non-pharmaceutical interventions (NPIs), to contain the spread of coronavirus infection in the population. These strategies include government interventions related to the close of education system, induce social isolation and voluntary quarantine. This suggests the inclusion of other variables on the mathematical modeling in order to have better adjustments with real data and to furnish useful information for decision making.

In this regard, hospital infrastructure and the number of deaths seem to be essential points to be included on mathematical modeling. The literature presents some research efforts related to the dynamics of COVID-19 pandemic progress considering different scenarios of the NPIs for reducing transmission of the virus, as well the hospital infrastructure necessary to take care of the infectious population (Canabarro *et al*., 2020; Ferguson *et al*., 2020; Lin *et al*., 2020; López & Rodó, 2020; Prem *et al*., 2020; Weissman *et al*., 2020).

This contribution proposes a mathematical model to describe the general propagation of the novel coronavirus. The idea is to use the SEIR framework including different novel aspects: removed population is represented by two populations – recovered and deaths; description of hospital infrastructure; and total population varies according to the number of deaths. Initially, a model verification is carried out considering infected population evolution of China, considered as a benchmark case due the availability of data for the entire cycle. COVID-19 dynamics is then investigated establishing different scenarios based on Brazilian data.

## 2. MATHEMATICAL MODEL

A frame-by-frame description of the COVID-19 dynamics can be represented by a set of differential equations of the form 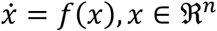, where *x* ∈ ℜ^*>n*^ represents a set of state variables that describe the phenomenon. The description of COVID-19 dynamics defines its propagation considering different kinds of populations. An interesting alternative for this aim is the susceptible-exposed-infectious-removed (SEIR) framework model. The populations are defined considering that *S* is the susceptible population, *E* is the exposed population, *I* is the infectious population, and two removed populations: recovered, *RD*, and death, *RD*, populations. Under this assumption, the total population is *N* = *S* + *E* + *I* + *R*_*C*_ + *R*_*D*_. Besides, the total population contains two classes: *D* is a public perception of risk regarding severe cases and deaths; and *C* represents the number of reported and non-reported cases. Another important observation is that population is reduced due to deaths, and therefore, *N* is reduced based on the increase of death population, *R*_*D*_, with a rate, 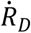.

Based on that, it is possible to write the following governing equations,

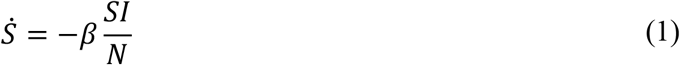

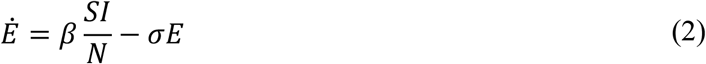

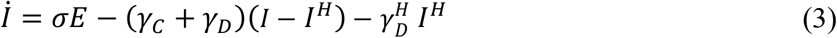

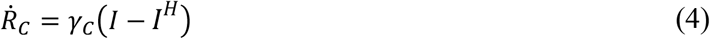

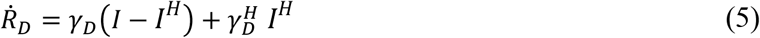

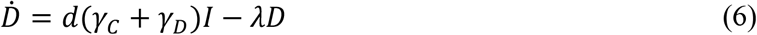

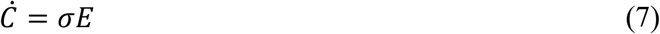

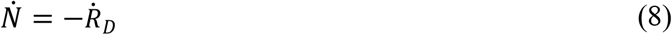

It should be pointed out that the model considers the effect of available hospital care for the infectious population. Therefore, it is defined a subpopulation of the infectious, *I*^*H*^, which represents the part of the infectious that needs hospital assistance but does not have access due to the lack of infrastructure. This is represented by function < > in order to represent the number of unavailable hospital assistance for the infectious population,

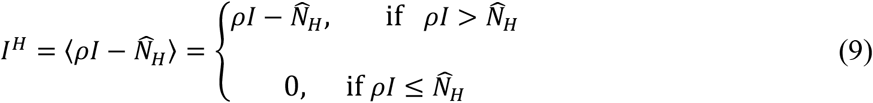

where *ρ* is the percentage of the population inside the group that needs hospital assistance and *NH* represents the number of available hospital infrastructure described with the aid of a step function as follows, and showed in Figure 1.

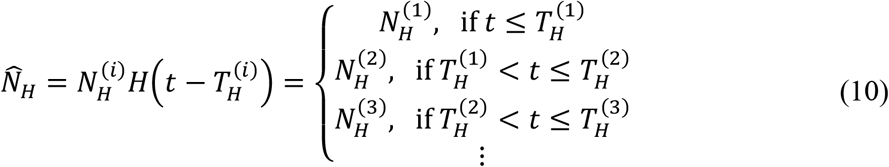

**Figure 1:**
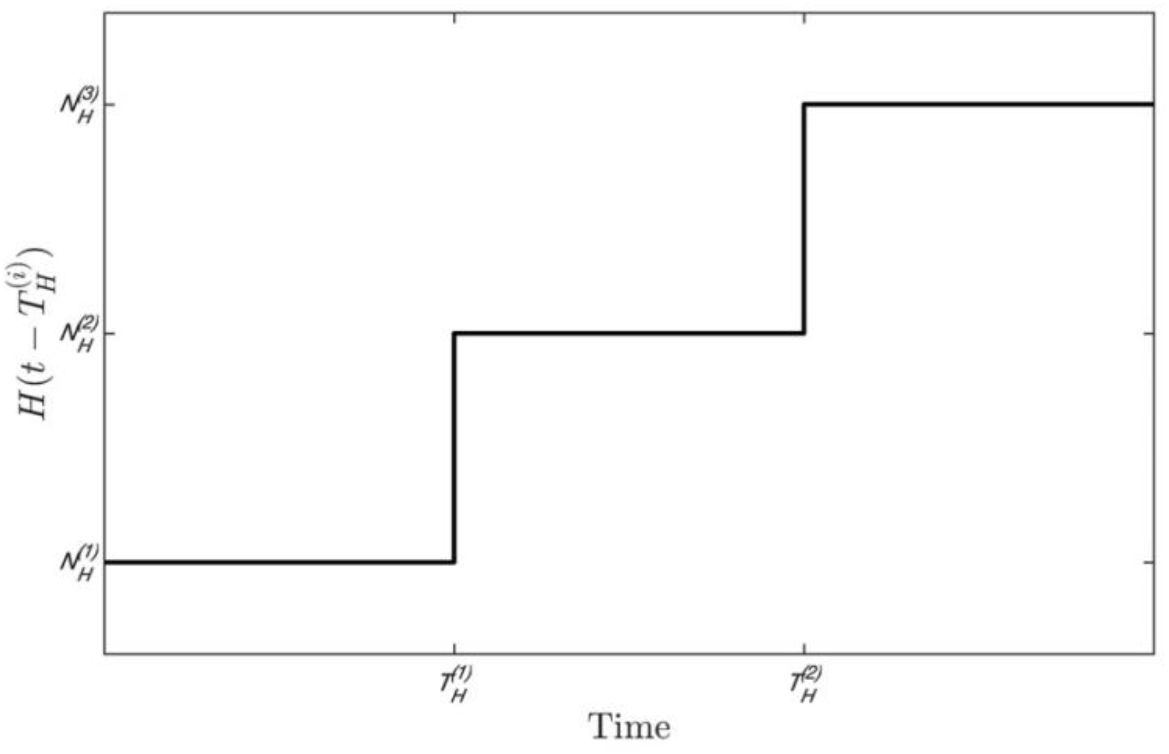
Step function employed to consider parameter variations through time.

The function *β* = *β*(*t*) represents the transmission rate that considers governmental action, represented by (1 − *α*); and the individual action, represented by the function *δ*. Therefore, the transmission rate is modeled as follows,

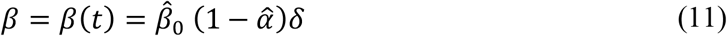

where 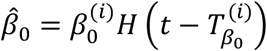 represents the nominal transmission rate and 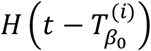 is a step function similar to the one previously defined. Using the same strategy, it is defined the governmental action as follows:

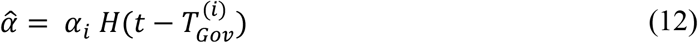

where different steps are considered defined by time instants 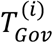.

In addition, individual action is represented by

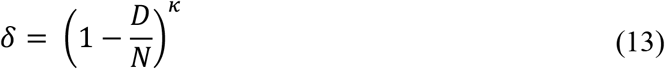

which the intensity of responses is defined by parameter *κ*. It should be pointed out that the different values of transmission rate are closely related to the social isolation. All these parameters need to be adjusted for each place, being essential for the COVID-19 description.

The following parameters are considered on the governing equations: *σ* is the mean latent period; *d* is the proportion of severe cases; *λ* is the mean duration of public reaction. Three parameters are adopted in order to describe the removed populations: *γ*_*C*_ is associated with the recovered population; *γ*_*D*_ is related to the death population; and 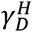 is related to the death population inside the group that needs hospital assistance, but due to system collapse, does not receive this assistance. The definition of the fatality rate is based on the relation between cumulative total deaths, *C*_*D*_, and the total cases, *C*_*I*_. Therefore, it is possible to use the following expression, considering a similar ratio between the associated rates for the removed population:

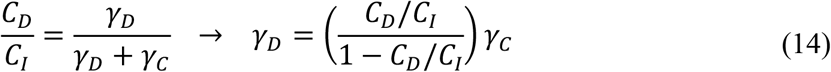

The recovered parameter, *γ*_*C*_, is defined based on the period necessary for the immune system. The death parameter, *γ*_*D*_, is defined from the expression presented in Eq. (14). The hospital parameter, 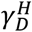 is defined from the relation of the part of the infected that needs hospital assistance.

In general, the parameter definitions depend on several issues, being a difficult task. In this regard, it should be pointed out that real data has spatial aspects that are not treated by this set of governing equations. Hence, this analysis is a kind of average behavior that needs a proper adjustment to match real data. The use of step functions to define some parameters allows a proper representation of different scenarios, including the transmission rate and hospital infrastructure. It is also important to observe that some researches concluded that undocumented novel coronavirus infections are critical for understanding the overall prevalence and pandemic potential of this disease. Li *et al*. (2020a) evaluated Wuhan situation and estimated that 86% of all infections were undocumented and that the transmission rate per person of undocumented infections was 55% of documented infections. This aspect makes the description even more complex. Table 1 presents parameters employed for all simulations. Other parameters are adjusted depending on the case.

**Table 1:** Model parameters for the simulations.

| Parameter | Description | Value |
| --- | --- | --- |
| $\gamma_D$ | Death rate | $\gamma_D = \left( \frac{C_D/C_I}{1 - R_D/C} \right) \gamma_C$ |
| $\gamma_D^H$ | Hospital assistance rate | 0.5 |
| $\rho$ | Percentage of the population inside the group that needs hospital assistance | 0.15 (15%) |
| $d$ | Perception of risk regarding severe cases and deaths | 0.2 |
| $\lambda^{-1}$ | Mean duration of public reaction | 11.2 days |
| $\kappa$ | Transmission rate modification parameter associated with individual actions | 1117.3 |

Numerical simulations are performed considering the fourth-order Runge-Kutta method. A convergence analysis is developed for the presented cases. The next sections treat the COVID-19 dynamics considering two different objectives. Initially, the next section performed a model verification using information from the process experienced by China. Afterward, the subsequent section evaluates different scenarios for the Brazilian case, using the parameters adjusted for the verification cases.

## 3. MODEL VERIFICATION

A model verification is carried out using information available on Worldometer (2020). It should be pointed out again that the analysis is based on average populations, assumed to have spatial homogeneous distribution. China data is considered as the benchmark case due to the large amount of information since it is the first case in the world and can be used to gather important information to support the predictions for other countries. This analysis is employed to calibrate the model parameters, evaluating its correspondence with real data. Table 2 presents additional parameters employed for the simulations. They are based on the information of the Lin *et al*. (2020) that, in turn, is based on other references as He *et al*. (2010) and Breto *et al*. (2009). Furthermore, an average value of the fatality rate 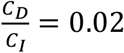 is observed from China actual data. It is important to highlight that this value is calculated with the available data and the existence of unreported cases can substantially change this number. For more details, see other citations referenced therein.

**Table 2:** Model parameters for China.

| Parameter | Description | Value |
| --- | --- | --- |
| $\beta_0$ | Nominal transmission rate | 0.514 |
| $\sigma^{-1}$ | Mean latent period | 3 days |
| $\gamma_C^{-1}$ | Mean recovered period | 5 days |
| $\gamma_D$ | Death rate | $4.082 \times 10^{-3}$ |
| $\alpha_i$ | Transmission rate modification parameter associated with governmental actions | [0, 0.4239, 0.8478] |
| $T_{Gov}^{(i)}$ | Transmission rate modification parameter associated with governmental actions | [0, 13, 20] days |

Parameters presented in Tables 1 and 2 are employed for simulations with a population of *N* = 1.43 billion and an initial state with 554 infected persons (*I*_0_ = 554), relative to January 22, 2020. In addition, susceptible population initial condition is assumed to be *S*_0_ = 0.9*N*. Another information needed for the model is the number exposed persons for each infected person. It is assumed that each infected person has the potential to expose 20 persons, *E*_0_ = 20*I*_0_.

Hospital infrastructure is considered to be without any restriction, which means that all the population that needs assistance is assisted and therefore, population *I*^*H*^ = 0 for all time during the whole simulation. Figure 2a presents infected and cumulative deaths populations showing a good agreement between simulation and real data obtained from Worldometer (2020). Figure 2b presents the evolution of other variables of the model. Note that, due to chronological issues, the whole cycle is observed on Chinese data. Based on that, it is possible to say that the model is capable to describe the whole cycle of COVID-19 dynamics.

**Figure 2:**
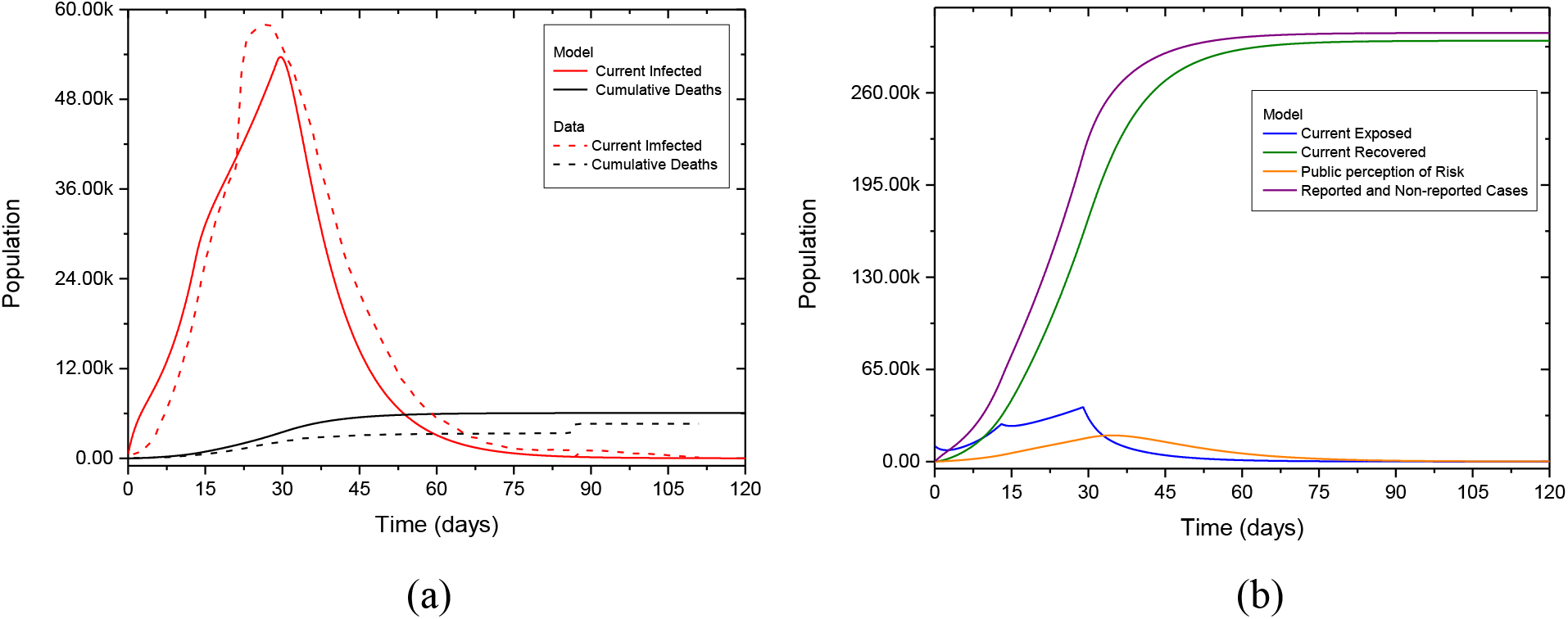
Model verification based on China actual data: (*a*) comparison between numerical results and data for infected and cumulative death population through time and (*b*) other model variables evolution.

## 4. BRAZILIAN SCENARIOS

This section has the objective to investigate different scenarios related to COVID-19 dynamics in Brazil. All simulations consider a population of *N* = 209.3 million and an initial state with 1 infected (*I*_0_ = 1) and 250 exposed persons (*E*0 = 250), relative to February 25, 2020. Parameters listed in Tables 1 and Table 3 are employed in all the simulations (Lyra *et al*., 2020; Ferguson *et al*., 2020). An average value of fatality rate 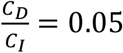 is observed for Brazil actual data. Values adopted for *α*_*i*_ consider three moments associated with governmental action (0, 15 and 45 days), representing the effect of two waves that occurred from the moment of the first infected person was identified and the present moment when the paper is being written. In the first wave, which occurred after 15 days, some regions of the country implemented actions of social isolation, such as closing the schools/universities and adopting remote work. One month after, there was a relaxation of the social isolation, which has been maintained until May, 2020.

**Table 3:** Model parameters for Brazil.

| Parameter | Description | Value |
| --- | --- | --- |
| $\beta_0$ | Nominal transmission rate | 1.020 |
| $\sigma^{-1}$ | Mean latent period | 5.2 days |
| $\gamma_C^{-1}$ | Mean recovered period | 2.9 days |
| $\gamma_D$ | Death rate | $1.815 \times 10^{-2}$ |
| $\alpha_i$ | Transmission rate modification parameter associated to governmental actions | [0, 0.40, 0.30] |
| $T_{Gov}^{(i)}$ | Transmission rate modification parameter associated to governmental actions | [0, 15, 45] days |

Figure 3a presents the infected population and cumulative deaths evolution obtained from numerical simulations and real data obtained from Worldometer (2020), showing that the same trend of the other cases is followed, being enough to have a general scenario. Figure 3b presents the evolution of other variables of the model. It should be highlighted that Brazilian outbreak is in the beginning, with information that is not enough for a better calibration.

**Figure 3:**
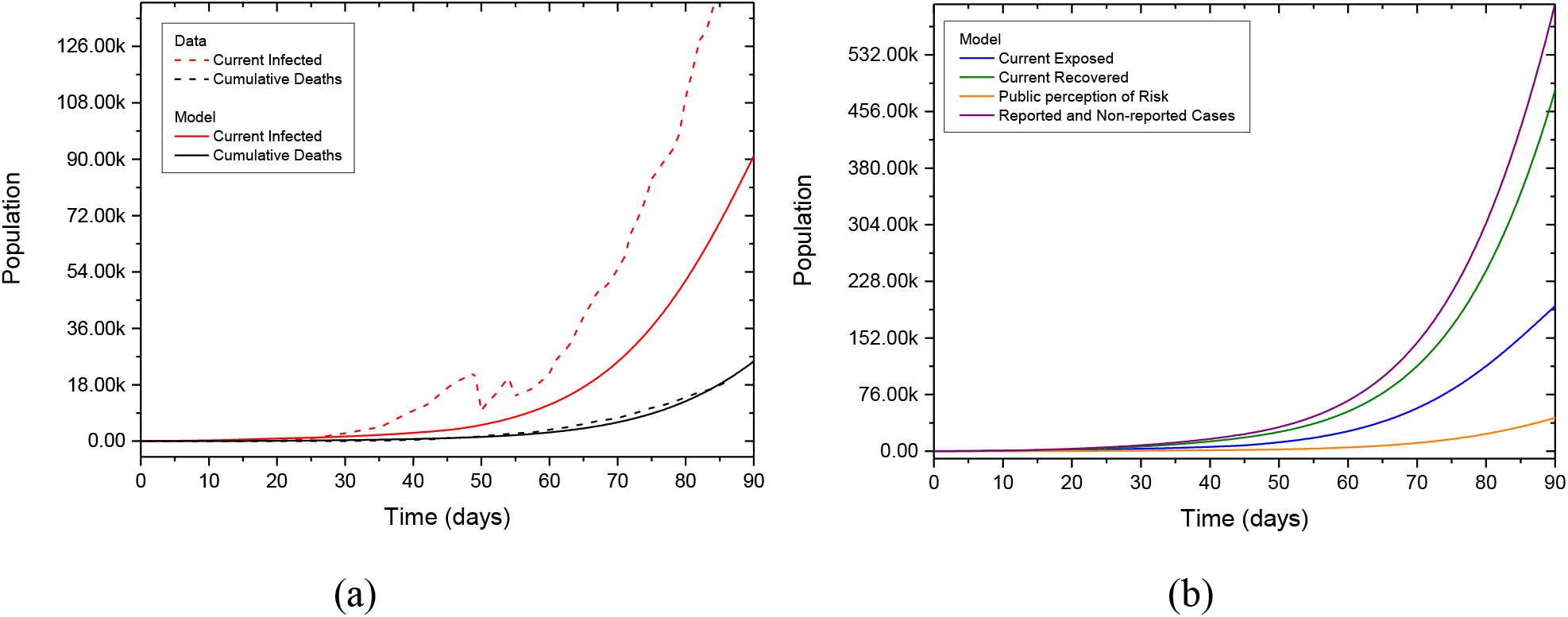
Model verification based on Brazil actual data: (*a*) comparison between numerical results and data for infected and cumulative death population through time and (*b*) other model variables evolution.

Different scenarios are now investigated considering a period of time until the end of 2020. Brazilian governmental action has the characteristic to be without a central coordination that makes the social isolation a polemic point, different from the great majority of the world. Based on that characteristic, it is important to present simulations showing distinct scenarios. Figure 4 presents different scenarios defined by the transmission rate changes, represented by governmental actions associated with different values of the parameter *α*, showing the evolution of all populations involved on COVID-19 dynamics. It is assumed an unlimited hospital infrastructure, which means that all the population that needs assistance is assisted. Calibrated values are maintained for the period of the first 90 days, defined from the available data for the present moment when the paper is being written. From this point forward, the future is predicted considering different values of *α*, which characterizes distinct scenarios. The following parameter values are adopted: 0.00, 0.30, 0.40, 0.50, 0.60, 0.70, 0.80, and 0.90. These scenarios represent different conditions associated with fixed government actions adopted at the end of the first 90 days that are maintained until the end of the year. It should be noticed that the curves have dramatic different values and therefore, the number of infected and deaths presents huge differences. There is a huge reduction of both numbers with the increase of social isolation. In addition, there is an important qualitative change related to the infectious population. The social isolation produces an infectious dynamics with a peak followed by a decrease to small numbers, called peak-vanish case, as showed in Figure 4b for *α* between 0.60–0.90. On the other hand, the lack of social isolation produces a curve with a plateau characteristic observed in Figure 4b for *α* between 0.00–0.50, which means that the critical period is spread over the time.

**Figure 4:**
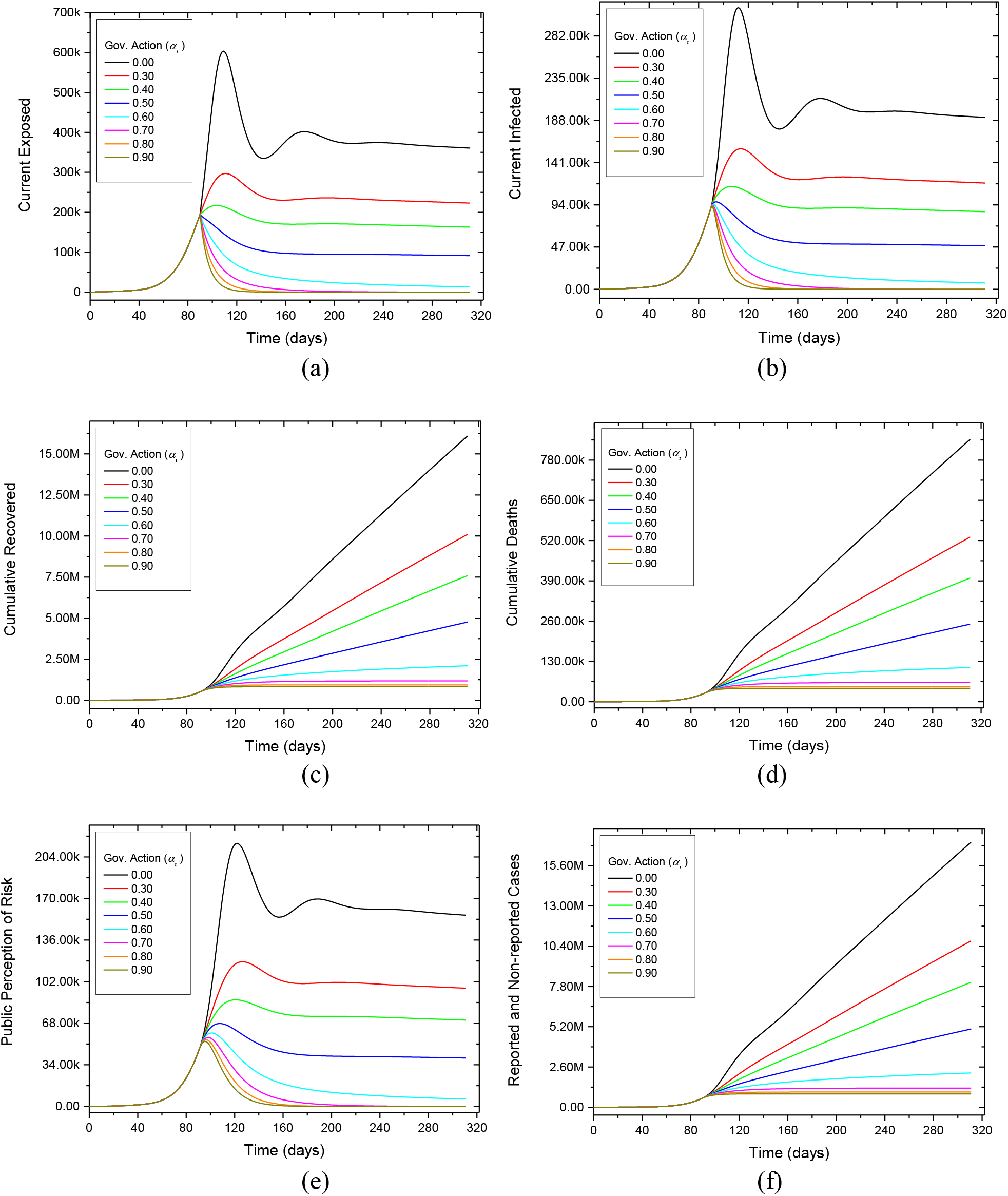
Population dynamics considering different governmental actions expressed by the social isolation: (*a*) current exposed, (*b*) current infected, (*c*) cumulative recovered, (*d*) cumulative deaths, (*e*) public perception risk and (*f*) reported and nom-reported cases. Hospital infrastructure without any restriction

Figure 5 presents detailed views of the dynamics of populations of current infected and cumulative deaths highlighting some characteristic behaviors. Once again, it is evident that the number of involved populations is dramatically different for each kind of governmental action. The difference of the two possible behaviors, characterized by the peak-vanish and plateau behaviors for the current infected populations, is also observed.

**Figure 5:**
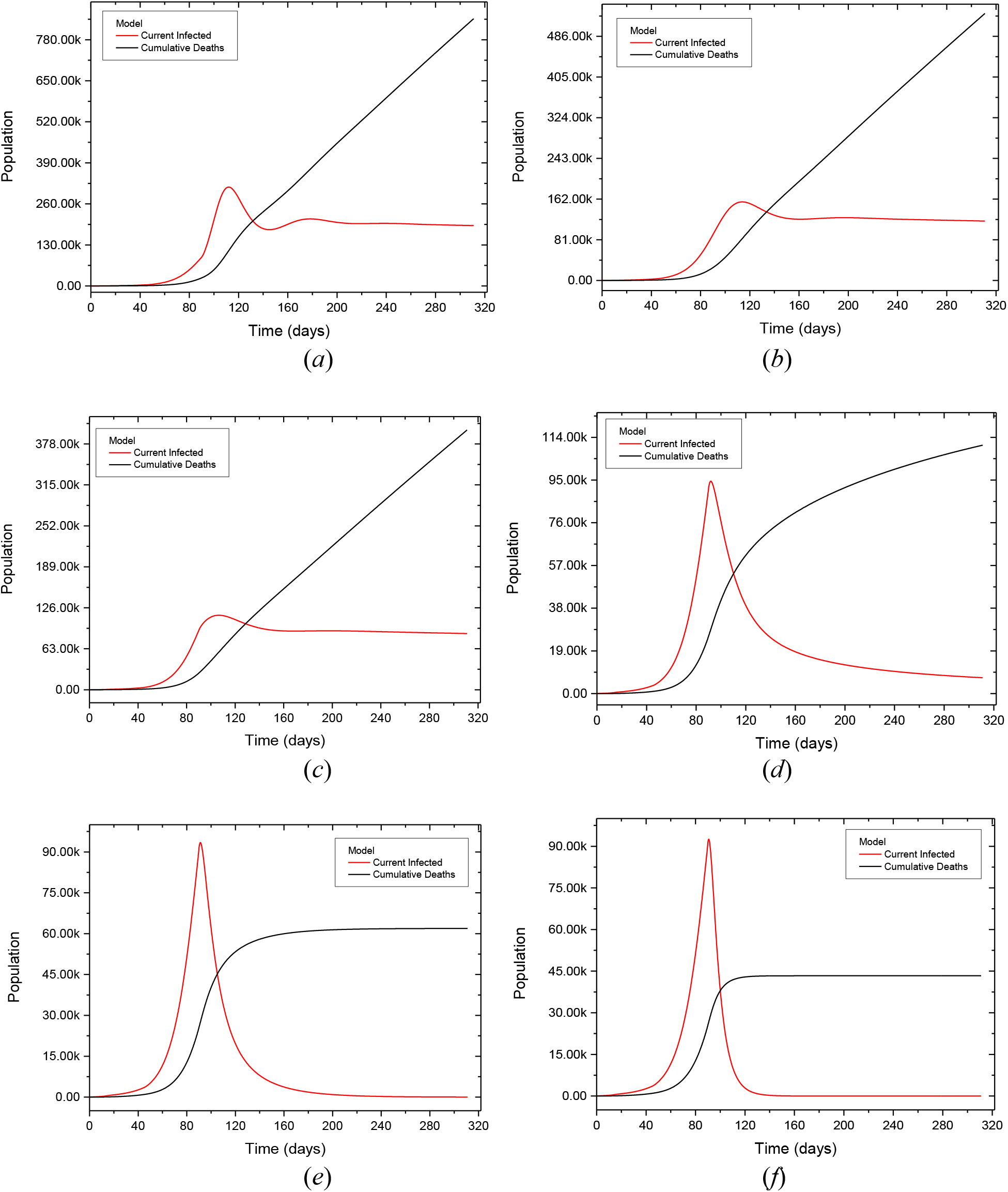
Population evolutions considering different transmission rates, altered by governmental actions: (*a*) *α* = 0.00, (*b*) *α* = 0.30, (*c*) *α* = 0.40, (*d*) *α* = 0.60, (*e*) *α* = 0.70, (*f*) *α* = 0.90.

Based on these scenarios, it should be pointed out that different governmental actions related to social isolation effect, result in dramatically different numbers of infected and cumulative deaths. Table 5 summarizes the results showing that a worst scenario of 846,833 deaths is in huge contrast with the best scenario of 43,331 deaths. This comparison clearly indicates that a more appropriate approach can result in a huge number of preserved lives.

**Table 4:**
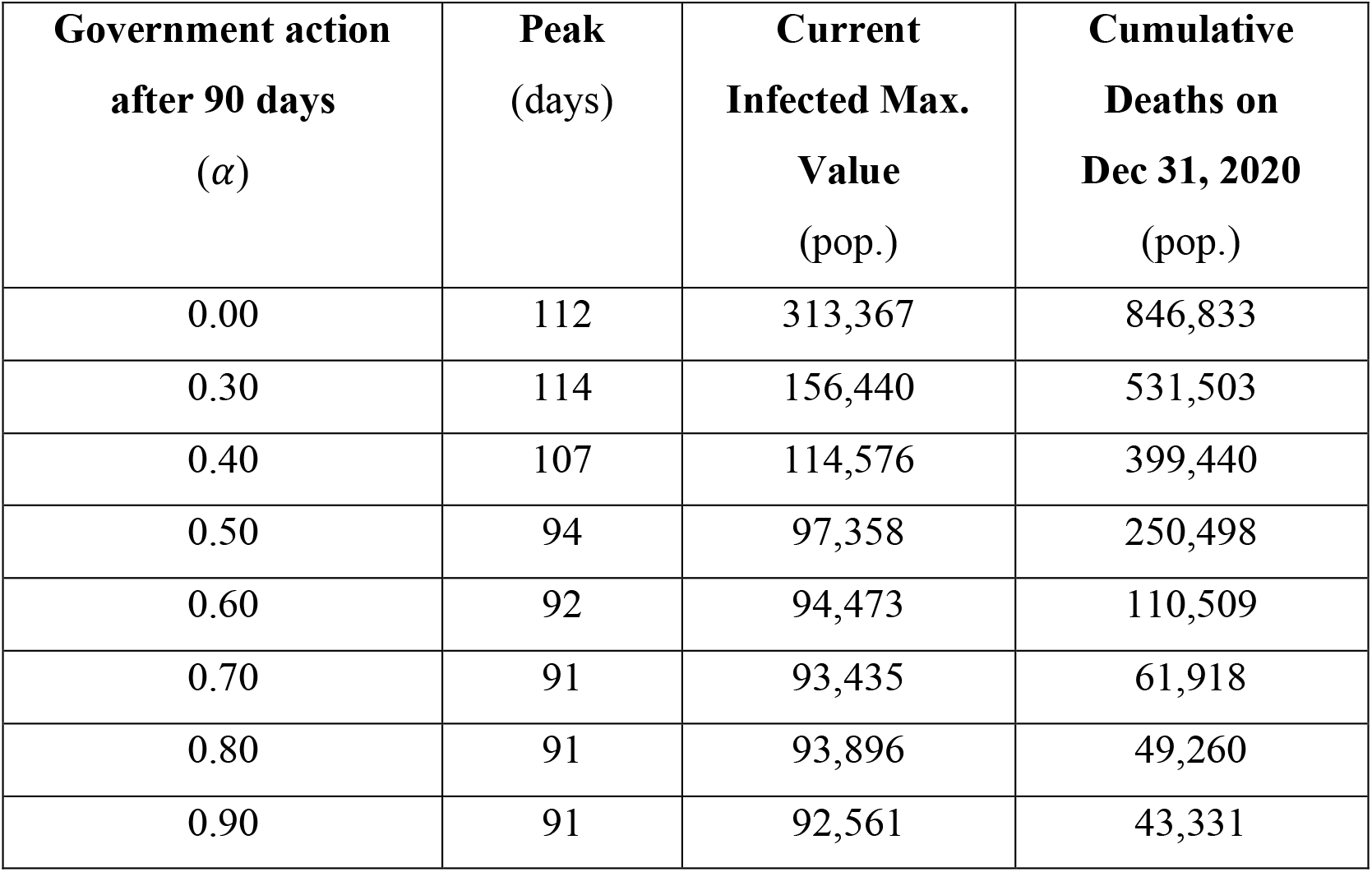
Infected and cumulative deaths predicted considering different government actions. Hospital infrastructure without any restriction.

One of the most relevant point related to COVID-19 evolution is the hospital infrastructure. Based on that, different scenarios are now of concern estimating distinct hospital infrastructure levels. The constraints are difficult to be quantified since it is not only the number of hospital beds available, but medical staff, drug availability and medical equipment are also necessary to define the hospital infrastructure number. The absence of this infrastructure increases the number of deaths since the population that needs assistance does not receive it. The specific infrastructure for this population is represented in the model by the total number of available Intensive Care Units (ICUs), designed by *NH*. Data accessed from the Brazilian Ministry of Health (2020a, 2020b) shows that Brazil has close to 40,000 ICUs. Nevertheless, only 13,939 are eligible for the treatment of patients with COVID-19. Numerical simulations are carried out considering this value of *N_H_*. It is important to highlight that there is a non-homogeneous geographic distribution in Brazil, with ratio of 9 and 21 ICUs beds per 100,000 inhabitants in the north and southeast regions of the country, respectively (Castro *et al*., 2020). In addition, some hospitals suffer with a lack of health professionals to assist patients with COVID-19. This condition can make the lack of ICUs even more critical.

Figure 6 presents the evolution of all populations involved on COVID-19 dynamics considering different scenarios defined for the distinct governmental actions treated on the previous simulations. Figure 7 shows the evolution of the population that needs hospital assistance and does not receive it. In general, it is noticeable the same qualitative behavior observed for the populations in the previous simulations for all cases, but two important points should be observed: the size of the populations is totally different, which means that the infected populations and deaths are completed different; the other important point to be observed is related to the hospital infrastructure. Note that there is a dramatic difference in terms of necessary hospital infrastructure, either for the number of hospital space or the spread over the time. The decrease of the social isolation is associated with the increase of the infected and death populations. In addition, infectious population presents an important plateau behavior that is related to an increase of deaths. Once again, it can be observed that different governmental actions result in dramatically different numbers of infected and cumulative deaths.

**Figure 6:**
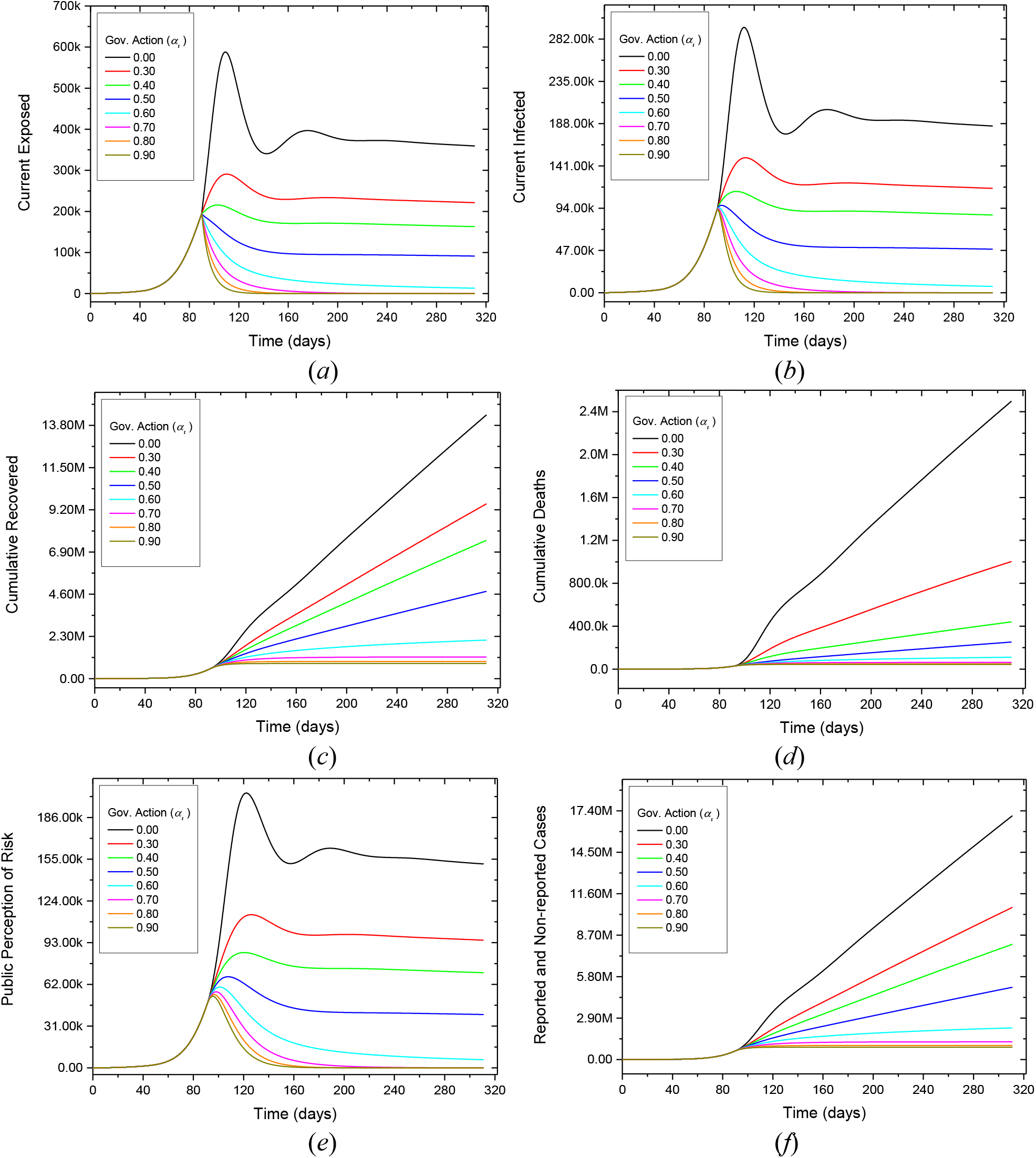
Population dynamics considering different governmental actions expressed by the social isolation: (*a*) current exposed, (*b*) current infected, (*c*) cumulative recovered, (*d*) cumulative deaths, (*e*) public perception risk and (*f*) reported and non-reported cases.

**Figure 7:**
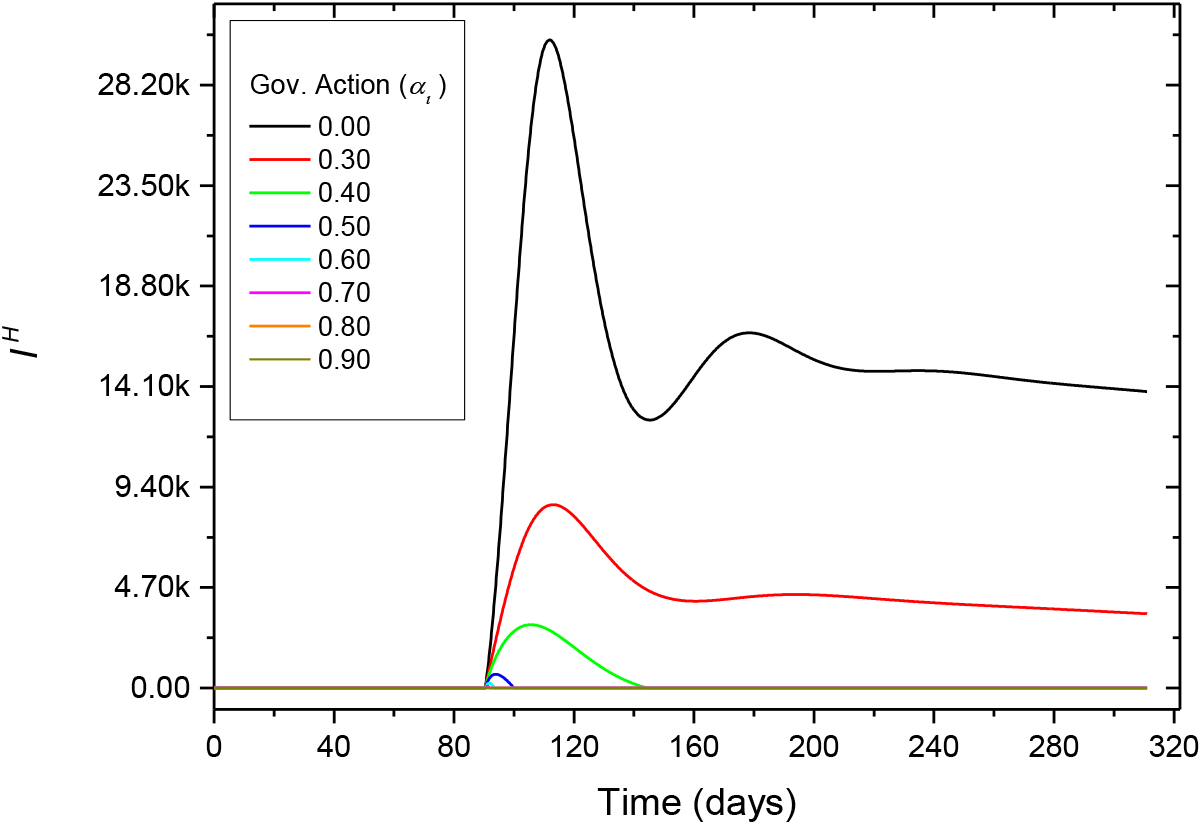
Population dynamics considering different governmental actions expressed by the social isolation. Infectious that needs hospital assistance but does not have access due to the lack of infrastructure.

Figure 8 highlights some characteristic behaviors of the dynamics. The left panel shows the populations of current infected and cumulative deaths, whereas the right panel shows the population that needs hospital assistance and does not receive it. As for the previous cases, the number of involved populations is dramatically different for each kind of governmental action, and two possible behaviors, characterized by the peak-vanish and plateau behaviors for the current infected populations, is also observed.

Table 5 summarizes the results showing a worst scenario of 2.5 million deaths (close to 1.2% of the Brazilian population) and a best scenario of 43,331 deaths, an even more dramatic difference when the hospital infrastructure is incorporated into the analysis. Results show that the limitations of the hospital infrastructure cause more than 30,000 infected individuals inside the group that needs hospital assistance may be left without access, a condition associated with a high fatality rate.

**Figure 8:**
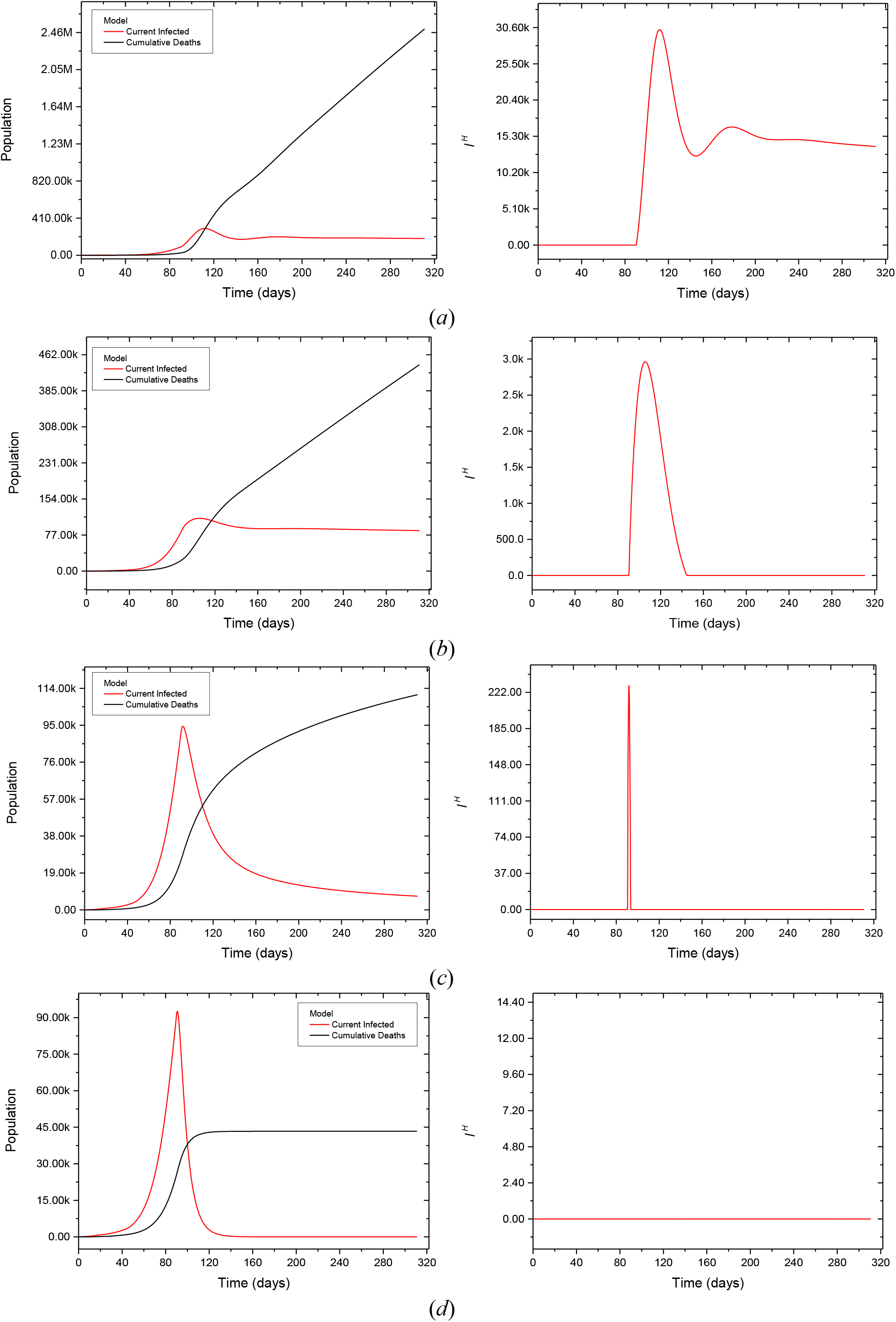
Population evolutions considering different transmission rates, altered by governmental actions: (*a*) *α* = 0.00, (*b*) *α* = 0.40, (*c*) *α* = 0.60, (*d*) *α* = 0.90.

**Table 5:** Infected and cumulative deaths predicted considering different government actions.

| **Government Intervention after 90 days ($\alpha$)** | **Current Infected - Peak (days)** | **Current Infected - Max. Value (pop.)** | **Cumulative Deaths on Dec 31, 2020 (pop.)** | **$I^H$ - Lack of Hospital Assistance (pop.)** |
| --- | --- | --- | --- | --- |
| 0.00 | 112 | 294,996 | 2,498,629 | 30,310 |
| 0.30 | 113 | 150,090 | 1,003,322 | 8,575 |
| 0.40 | 106 | 112,670 | 440,749 | 2,962 |
| 0.50 | 94 | 97,210 | 252,369 | 643 |
| 0.60 | 92 | 94,452 | 110,715 | 229 |
| 0.70 | 91 | 93,432 | 61,948 | 76 |
| 0.80 | 91 | 92,896 | 49,260 | 0 |
| 0.90 | 91 | 92,561 | 43,331 | 0 |

Figure 9 shows a comparison between numerical results considering two situations relative to the specific hospital infrastructure required to deal with the COVID-19. The first set of results (Unlimited Hosp. Infrastructure) considers an ideal condition where there is no restriction of the hospital infrastructure to assist part of the infectious that needs hospital assistance, whereas the second one (Limited Hosp. Infrastructure) considers situations where restrictions are defined by the total number of available Intensive Care Units (ICUs). Numerical results confirm that the absence of this infrastructure largely increases the number of deaths since the population that needs assistance does not receive it. This increase the deaths from 846,833 to 2,498,629 (for *α* = 0.00 after 90 days) is an emblematic situation associated with an increase of more than 200%.

**Figure 9:**
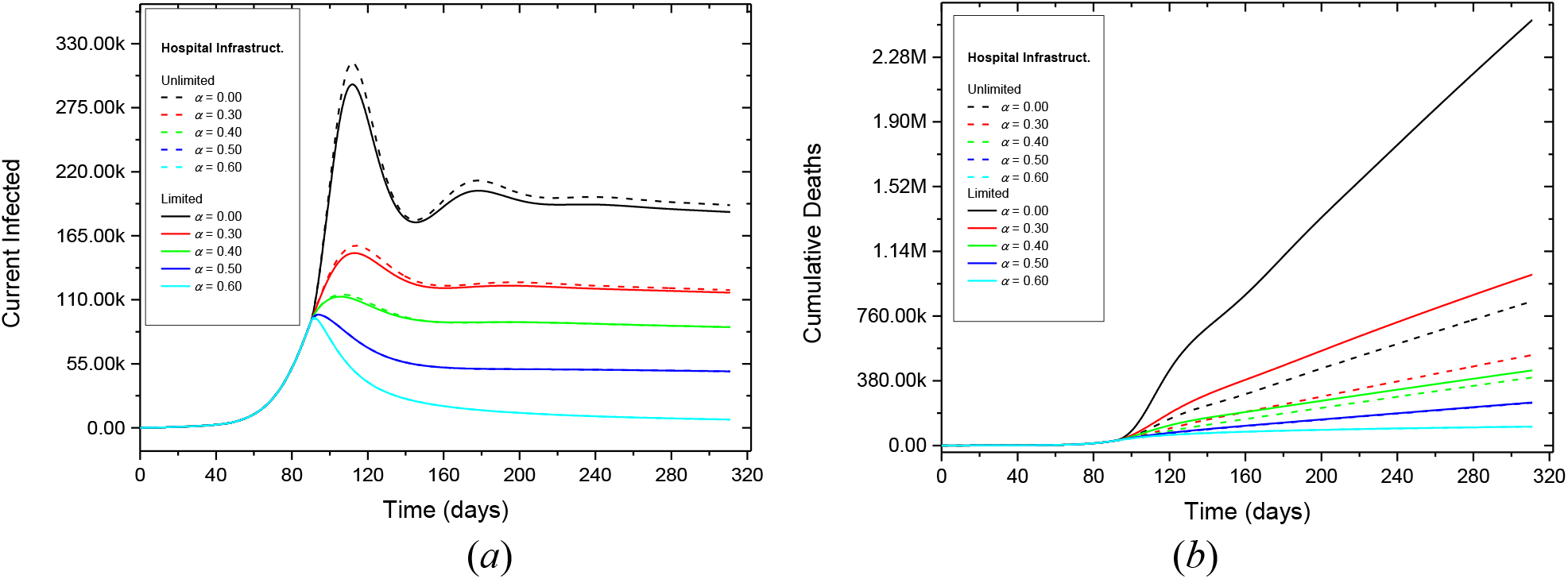
Influence of the hospital infrastructure limitations on the population dynamics considering different governmental actions expressed by the social isolation: (*a*) current infected, (*b*) cumulative deaths.

Several possibilities for the social isolation have been discussed worldwide. One promising scenario involves the combination of triggering hardening/softening actions (Fergurson *et al*., 2020). Therefore, different scenarios are now evaluated considering several implementation approaches for governmental actions. Table 6 presents cases where the previous adopted values of *α* for the interventions prior to 90 days are maintained and future actions are implemented considering a combination of hardening/softening governmental actions. Results from the previous analysis show that a minimum value of *α* = 0.70 must be adopted at 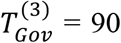 days to maintain the number of deaths bellow 100,000. Therefore, this minimum value is adopted for the analysis.

**Table 6:**
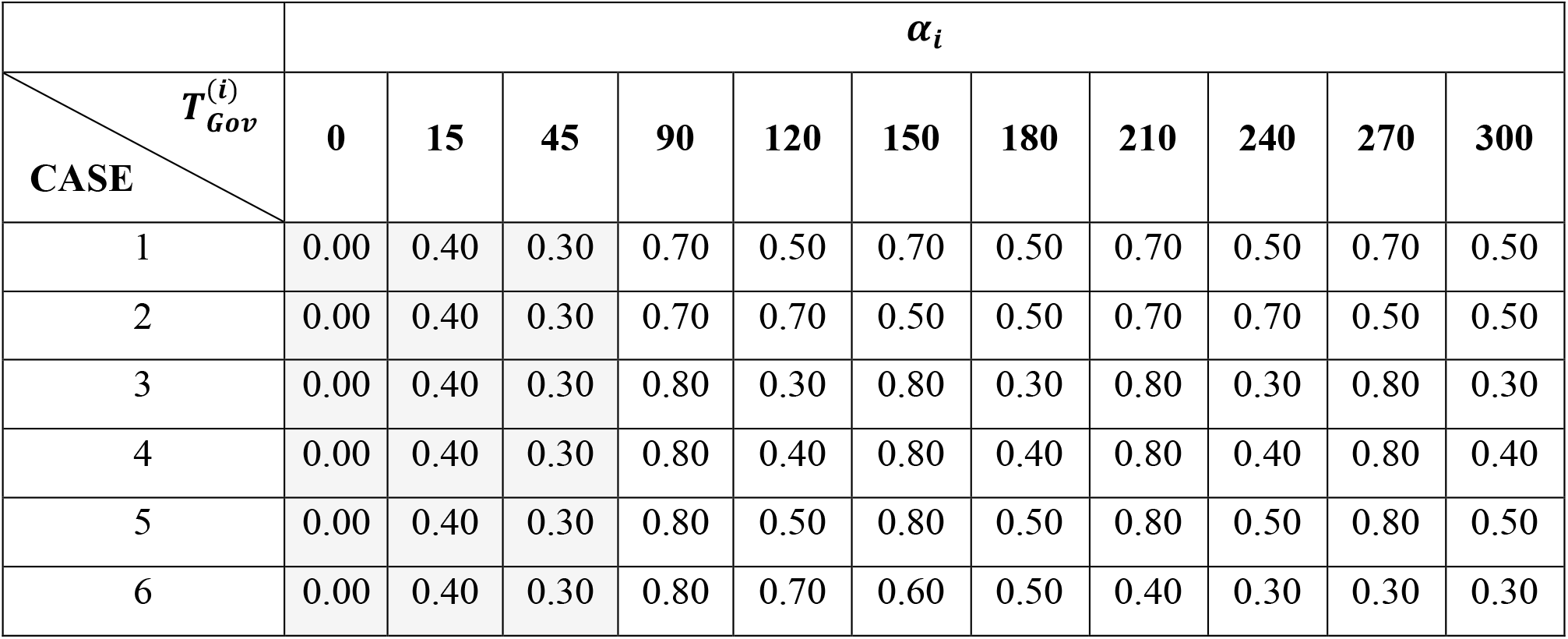
Different scenarios considering several implementation approaches of governmental actions. Interventions prior to 90 days preserved.

Figure 10 shows the population evolutions considering Cases 1–6. Cases 1–5 are associated with cyclic governmental actions. These actions result in multiple subsequent infection waves. Case 6 represents a condition where a governmental hardening action is implemented after 90 days, followed by a progressive softening action. For this situation, the increase of the infectious population during a second wave, together with the limitations of the available hospital infrastructure, result in more than 8,000 infected individuals within the group who needs hospital assistance. This contribute for a larger number of deaths that could be avoided. These scenarios show that social isolation combined with proper hospital infrastructure can drastically reduce the number of deaths.

**Figure 10:**
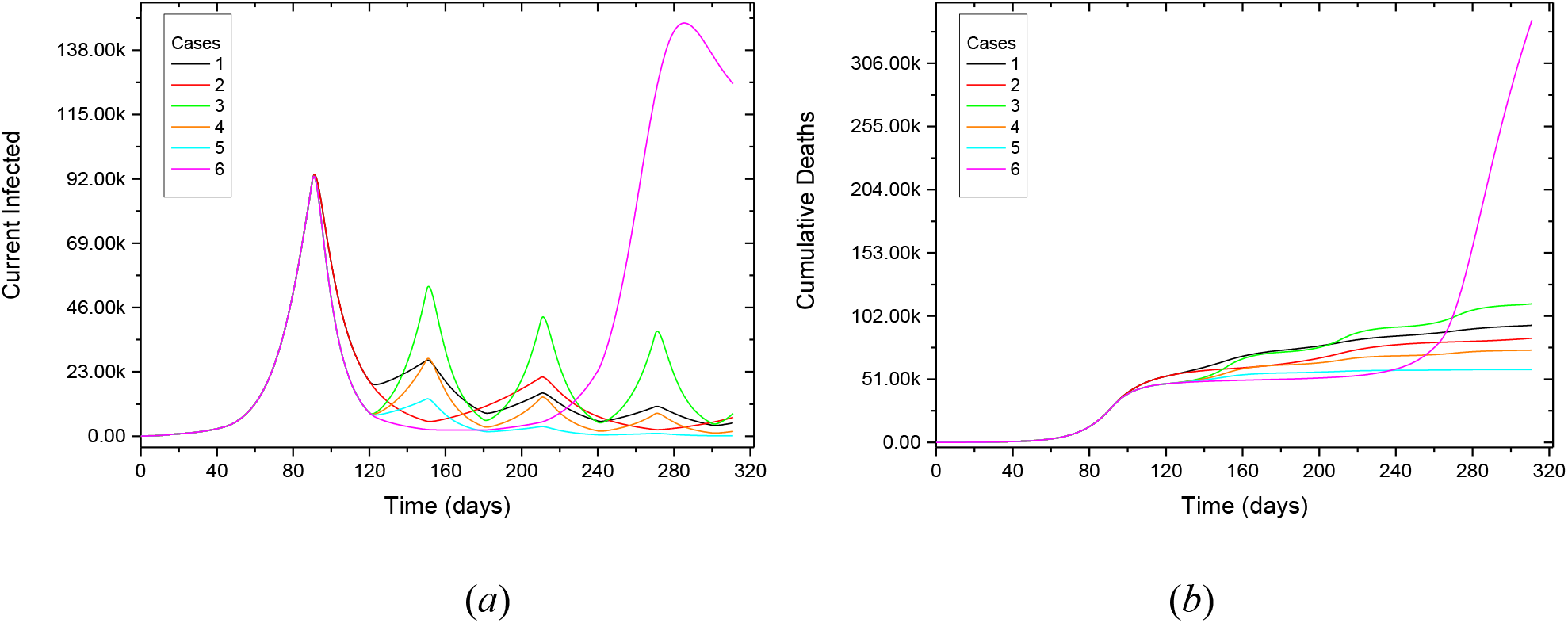
Population dynamics considering different governmental actions expressed by the social isolation: (*a*) current infected, (*b*) cumulative deaths.

Figure 11 highlights some characteristic behaviors of the population dynamics of the current infected and cumulative deaths for some cases associated with cyclic governmental actions, whereas Figure 12 presents results for a governmental hardening action followed by a progressive softening action. The left panel shows the populations of the current infected and cumulative deaths, whereas the right panel shows the population that needs hospital assistance and does not receive it. Results show multi peak behavior followed by vanish or plateau behaviors for the current infected populations.

**Figure 11:**
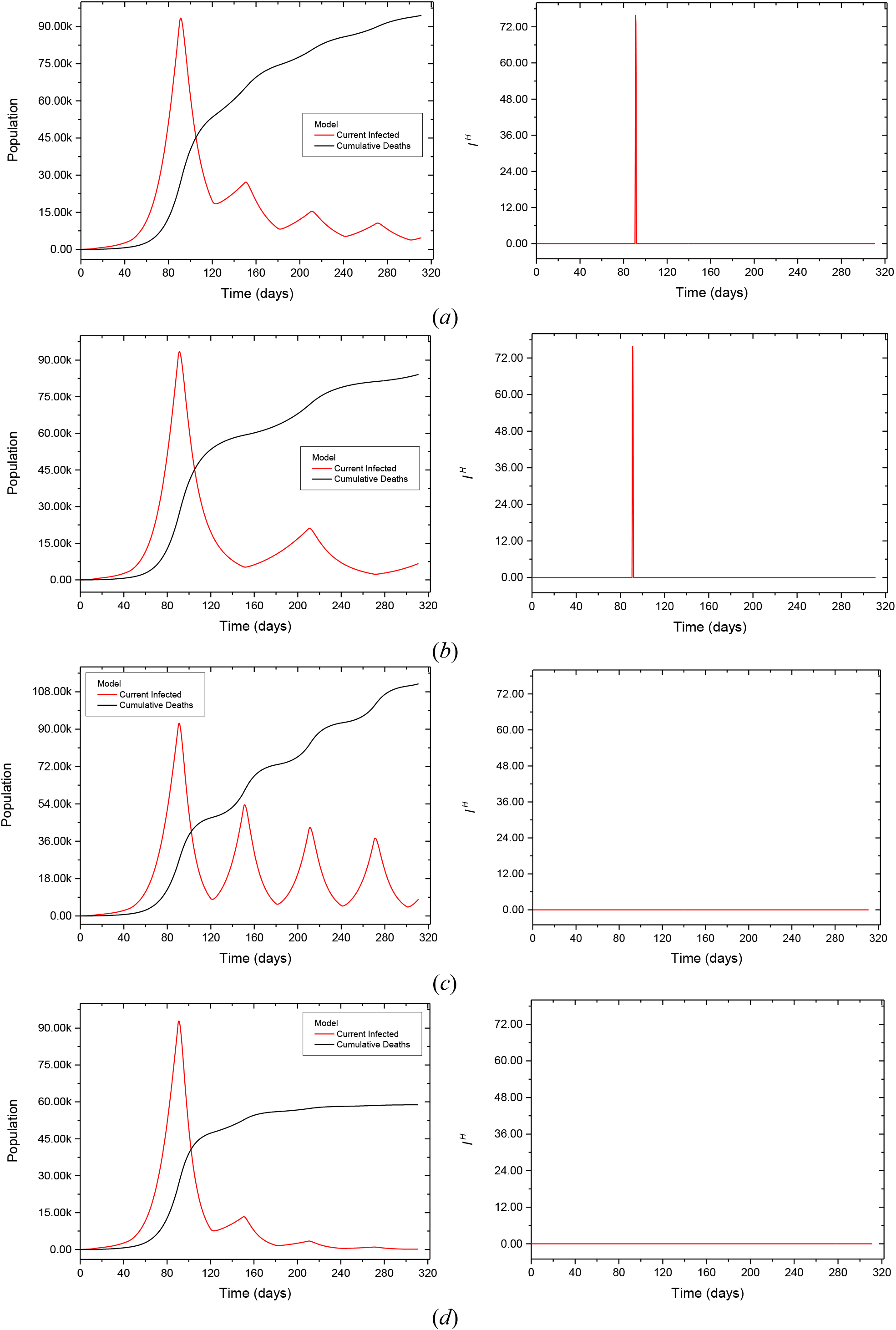
Population evolutions considering different transmission rates, altered by cyclic governmental actions: (*a*) Case 1, (*b*) Case 2, (*c*) Case 3, and (*e*) Case 5.

**Figure 12:**
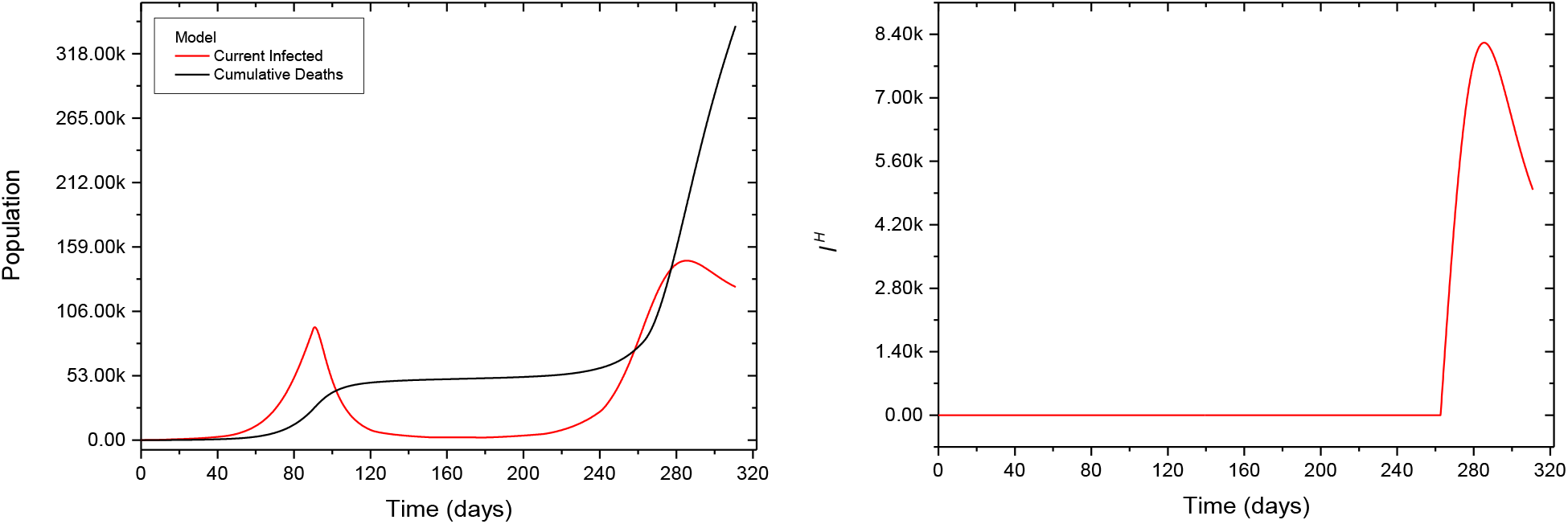
Population evolutions considering different transmission rates, altered by a governmental hardening action followed by a progressive softening action: Case 6. Interventions prior to 90 days preserved.

Table 7 summarizes these results, showing a worst scenario of 340,934 deaths and a best scenario of 58,831 deaths, indicating that a reduction of the number of deaths is possible with a proper isolation strategy controlled by governmental action.

**Table 7:** Infected and cumulative deaths predicted considering different government actions associated to cyclic actions and hardening action followed by a progressive softening action.

| **Government Intervention after 90 days ($\alpha$)** | **Current Infected - Peak (days)** | **Current Infected - Max. Value (pop.)** | **Cumulative Deaths on Dec 31, 2020 (pop.)** | **$I^H$ - Lack of Hospital Assistance (pop.)** |
| --- | --- | --- | --- | --- |
| Case 1 | 91 | 93,432 | 94,520 | 76 |
| Case 2 | 91 | 93,432 | 84,111 | 76 |
| Case 3 | 91 | 92,896 | 111,845 | 0 |
| Case 4 | 91 | 92,896 | 74,448 | 0 |
| Case 5 | 91 | 92,896 | 58,831 | 0 |
| Case 6 | 91 / 286(*) | 92,896 / 147,700(*) | 340,934 | 8,216 |

(\*) second wave peak

As a highly contagious disease, COVID-19 requires the implementation of governmental actions in the very begging. Previous analysis has shown the difficulties to control or reduce the number of infectious and deaths if nothing is done at early stages. With the objective to analyze the importance of the implementation of a rapid response, three approaches involving early government actions are considered: Constant; Progressive; and Cyclic. In the first one, a constant governmental action is adopted from the beginning of the intervention; the second considers a progressive reduction of the governmental action until a value of 0.50 is reached; the third one considers a cyclic variation between two levels of governmental action with a period of two months. For all the cases, the governmental action begins in the 15th day. Table 8 presents the cases description and Table 9 summarizes results for the three approaches. Due to the early actions taken, none of the 9 cases presented a condition of lack of hospital infrastructure for the part of the infectious population requiring specific assistance. Figure 13 presents the evolution of the current infected (left panel) and the cumulative deaths (right panel).

**Table 8:**
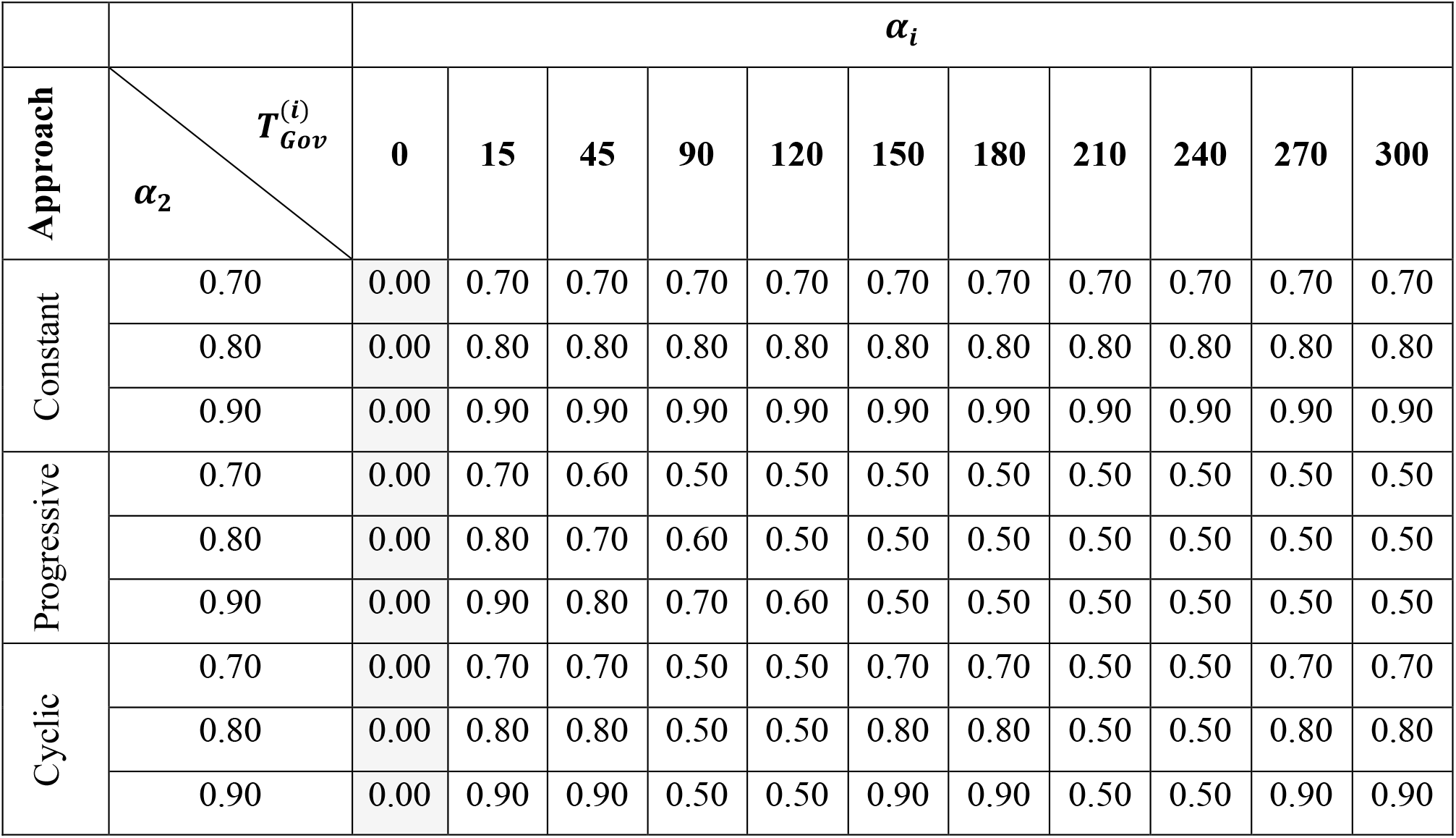
Different scenarios considering different early governmental actions approaches: constant, progressive and cyclic.

**Table 9:** Infected and cumulative deaths predicted considering different early governmental actions approaches: Constant, Progressive and Cyclic.

| **Approach** | **Government Intervention ($\alpha$)** | **Current Infected - Peak (days)** | **Current Infected - Max. Value (pop.)** | **Cumulative Deaths on Dec 31, 2020 (pop.)** |
| --- | --- | --- | --- | --- |
| Constant | 0.70 | 18 | 636 | 469 |
|  | 0.80 | 17 | 614 | 259 |
|  | 0.90 | 17 | 602 | 191 |
| Progressive | 0.70 | 18 / 311(*) | 635 / 51,386(*) | 51,386 |
|  | 0.80 | 17 / 311(*) | 614 / 11,969(*) | 11,969 |
|  | 0.90 | 17 | 602 | 196 |
| Cyclic | 0.70 | 18 | 636 | 1,188 |
|  | 0.80 | 17 | 614 | 277 |
|  | 0.90 | 17 | 602 | 191 |

(\*) no peak – value in the 311 day

**Figure 13:**
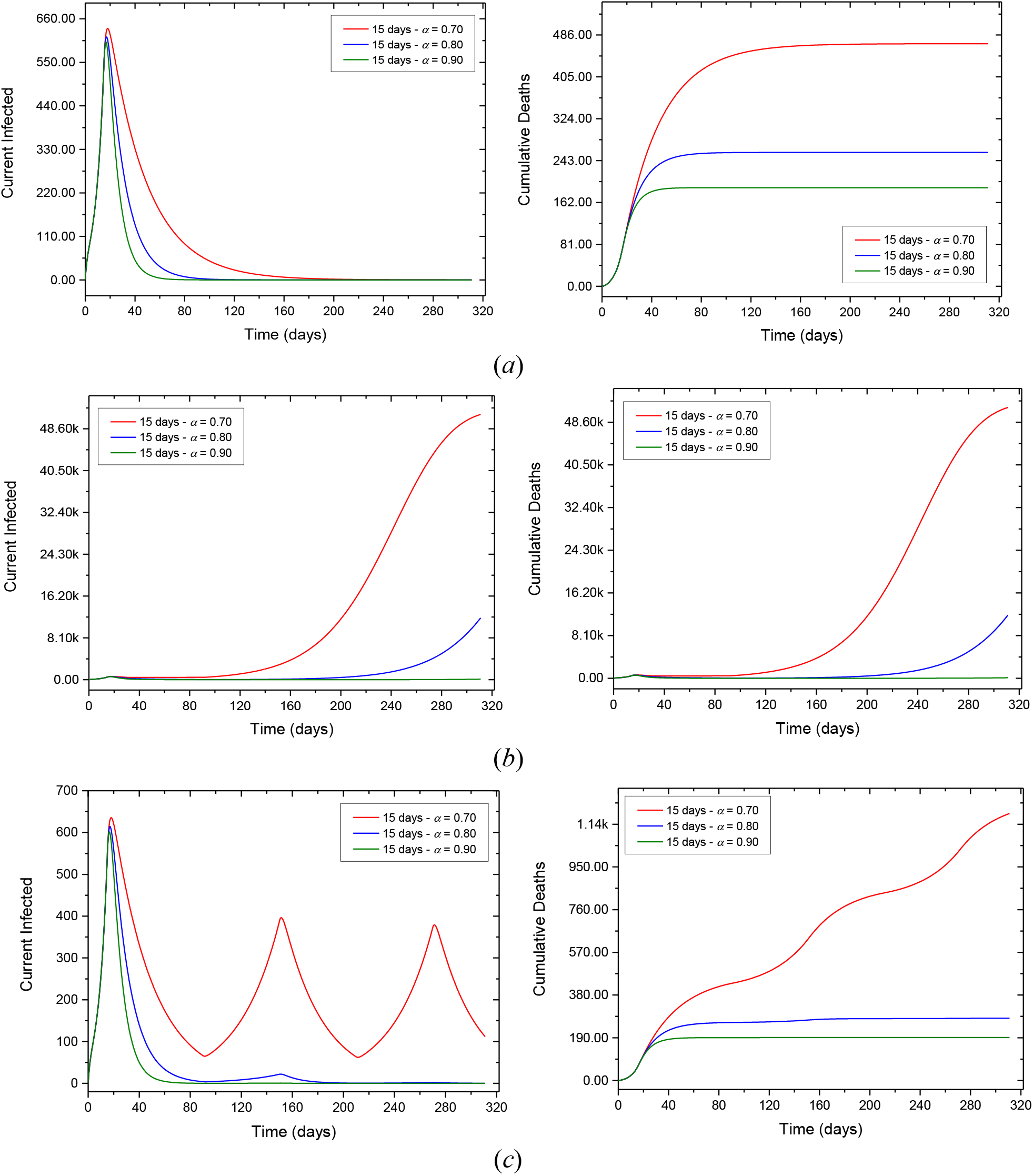
Evolution of the infected and cumulative deaths populations considering different early governmental actions approaches implemented at the beginning of the pandemic in Brazil: (*a*) Constant, (*b*) Progressive and (*c*) Cyclic. Early government intervention (prior to 90 days).

These results show that, in comparison with previous cases, the use of early governmental actions causes a smaller population of infectious, for which the cumulative deaths can be below 200. There is also no shortage of hospital infrastructure for the part of the infectious population requiring assistance. Overall, the Constant approach presents the best results associated with a smaller number of cumulative deaths at the end of the period, but with the cost of maintain a severe level of social isolation during a long period of time. The use of an initial *α* value of 0.90 results in a similar behavior for the three approaches, with a number of cumulative deaths lower than 200. For initial values of *α* of 0.70 and 0.80, the Progressive approach reveals the effect of a second wave (as in Case 6 of the previous analysis), resulting in a large number of deaths. A similar behavior is observed considering the Cyclic approach for *α* = 0.70, where many deaths are observed due to a second and a third infectious waves. However, for *α* = 0.80 the cyclic approach furnishes lower values of cumulative deaths, similar to the constant approach. Therefore, it can be an interesting alternative to replace the constant approach with the advantage of imposing a less severe governmental action and, therefore, a condition of less severe social isolation over the whole period.

It is important to highlight that, due to the strong sensitivity of the system nonlinear dynamics, small changes in conditions or control parameters can greatly affect the evolution of populations. Therefore, success in controlling the pandemic and reducing deaths depends on the adoption of approaches and mechanisms that allow monitoring the evolution of populations together with the rapid implementation of control procedures in the form of efficient government actions.

## 5. CONCLUSIONS

A mathematical model based on the susceptible-exposed-infectious-recovered framework is employed to describe the COVID-19 evolution. The proposed model considers removed populations composed by recovered and deaths populations. In addition, a population that needs hospital assistance is incorporated allowing the analysis of the lack of hospital infrastructure. A benchmark case is treated considering available data from China. Afterward, Brazilian case is analyzed. Initially, a verification case is performed with available data showing the capability of the model to describe real data. Afterward, different scenarios are of concern investigating governmental actions and hospital infrastructure, evaluating their evolution until the end of 2020. Numerical simulations clearly show that social isolation, guided by governmental and individual actions, are essential to reduce the infected populations and the total period of the crisis. Results based on actual data show that the number of deaths can vary from 40 thousand to 2.5 million depending on the social isolation level and the hospital infrastructure. In addition, early governmental actions are essential to ensure smaller population infectious and the absence of shortage of hospital infrastructure for the part of the infectious population requiring assistance, for which numerical simulations indicate a total number of deaths below 200. Therefore, simulations present differences from 200 to 2.5 million deaths, showing that a central coordination is essential to save a huge number of lives. Different qualitative behaviors can be expected depending on social isolation levels. On one hand, it is possible to observe a peak-vanish infectious curve representing a rapid dramatic crisis. On the other hand, COVID-19 dynamics can present a plateau behavior, spreading the crisis for a long period of time, being even more dramatic in terms of the number of deaths. Although the mathematical model can be improved in order to include more phenomenological information that can increase its capability to describe different scenarios, it should be pointed out that numerical simulations seem to be coherent with available data, being an important tool that can be useful for public health planning.

## Data Availability

NA

## 6. ACKNOWLEDGEMENTS

The authors would like to acknowledge the support of the Brazilian Research Agencies CNPq, CAPES and FAPERJ.

## Notes

### Competing Interest Statement

The authors have declared no competing interest.

